# Post-Discharge Exposure to Ambient Sulfur Dioxide is Associated with Increased Risk of Stroke Recurrence

**DOI:** 10.64898/2026.05.11.26352955

**Authors:** Keon-Joo Lee, Jeongeun Hwang, Seong-Eun Kim, Beom Joon Kim, Moon-Ku Han, Hyunsoo Kim, Joon-Tae Kim, Kang-Ho Choi, Kyu Sun Yum, Dong-Ick Shin, Jae-Kwan Cha, Dae-Hyun Kim, Dong-Seok Gwak, Dong-Eog Kim, Jong-Moo Park, Kyusik Kang, Soo Joo Lee, Jae Guk Kim, Minwoo Lee, Mi-Sun Oh, Kyung-Ho Yu, Hong-Kyun Park, Keun-Sik Hong, Yong-Jin Cho, Joong-Goo Kim, Jay Chol Choi, Tai Hwan Park, Sang-Soon Park, Jee-Hyun Kwon, Wook-Joo Kim, Doo Hyuk Kwon, Jun Lee, Kyungbok Lee, Jeong-Yoon Lee, Sung Il Sohn, Jeong-Ho Hong, Kwang-Yeol Park, Hae-Bong Jeong, Chulho Kim, Sang-Hwa Lee, Juneyoung Lee, Hee-Joon Bae, the CRCS-K-NIH investigators

**Affiliations:** Department of Neurology, Korea University College of Medicine, Korea University Guro Hospital, Seoul, Korea; Department of Medical IT Engineering, College of Software Convergence, Soonchunhyang University, Asan, Korea; Department of Neurology, Seoul National University College of Medicine, Seoul National University Bundang Hospital, Seongnam, Korea; Department of Neurology, Chonnam National University Hospital, Gwangju, Korea; Department of Neurology, Chungbuk National University & Hospital, Cheongju, Korea; Department of Neurology, Dong-A University Hospital, Busan, Korea; Department of Neurology, Dongguk University Ilsan Hospital, Goyang, Korea; Department of Neurology, Uijeongbu Eulji Hospital, Eulji University School of Medicine, Seoul, Korea; Department of Neurology, Nowon Eulji Medical Center, Eulji University School of Medicine, Seoul, Korea; Department of Neurology, Eulji University Hospital, Eulji University School of Medicine, Daejeon, Korea; Department of Neurology, Hallym University Sacred Heart Hospital, Anyang, Korea; Department of Neurology, Inje University Ilsan Paik Hospital, Goyang, Korea; Department of Neurology, Jeju National University Hospital, Jeju, Korea; Department of Neurology, Seoul Medical Center, Seoul, Korea; Department of Neurology, Ulsan University Hospital, Ulsan, Korea; Department of Neurology, Yeungnam University Medical Center, Daegu, Korea; Department of Neurology, Soonchunhyang University Seoul Hospital, Seoul, Korea; Department of Neurology, Keimyung University Dongsan Medical Center, Daegu, Korea; Department of Neurology, Chung-Ang University Hospital, Seoul, Korea; Department of Neurology, Hallym University Chuncheon Sacred Heart Hospital, Chuncheon, Korea; Department of Biostatistics, Korea University, Seoul, Korea

**Keywords:** sulfur dioxide, air pollution, ischemic stroke, stroke recurrence, secondary prevention, competing risk, marginal Cox model

## Abstract

**Background and Purpose:** Ambient air pollution is an established risk factor for incident stroke, but whether post-discharge pollutant exposure influences stroke recurrence remains unknown. We investigated the association between post-discharge exposure to six ambient air pollutants and stroke recurrence in patients with acute ischemic stroke.

**Methods:** We analyzed data from 27,346 patients in the CRCS-K-NIH nationwide multicenter registry of acute ischemic stroke patients (2014–2021) with confirmed ischemic stroke, residential address data, and matched air quality records. The primary exposure was the 3-month post-discharge average concentration of PM_10_, PM_2.5_, NO₂, SO₂, CO, and O₃, assessed at the district level using inverse-distance weighted interpolation. The primary outcome was stroke recurrence from 3 to 15 months post-discharge. Cause-specific Cox proportional hazards models accounting for the multilevel data structure were used, with all-cause mortality as a competing risk. Restricted cubic splines assessed nonlinear dose-response relationships.

**Results:** During follow-up (median 364.8 days), 765 patients experienced stroke recurrence and 471 died. Among the six pollutants, only SO₂ showed a statistically significant association with recurrence (P for overall association in the restricted cubic spline analysis = 0.024). A potential threshold was identified at approximately 8.2 ppb, above which recurrence risk increased progressively (P for non-linearity = 0.095). The association was numerically stronger among older adults (≥75 years; P for interaction = 0.051) and women (P for interaction = 0.062). The highest SO₂ concentrations were observed in harbor cities (Incheon, Ulsan, Busan), consistent with maritime shipping emissions. No significant associations were observed for the other five pollutants.

**Conclusions:** Elevated post-discharge SO₂ exposure is associated with increased stroke recurrence risk, particularly in harbor regions and among older adults and women. These findings support incorporating ambient air quality monitoring into secondary stroke prevention strategies.

## INTRODUCTION

Ambient air pollution is increasingly recognized as one of the leading modifiable environmental risk factors for cardiovascular and cerebrovascular diseases.^1,2^ Large-scale epidemiologic studies have estimated that nearly 20% of all cardiovascular mortality worldwide is attributable to air pollution,^2,3^ with fine particulate matter (PM_2.5_), nitrogen dioxide (NO₂), sulfur dioxide (SO₂), carbon monoxide (CO), and ozone (O₃) each demonstrating independent associations with incident ischemic stroke.^4–6^ These findings have driven international efforts to integrate air quality standards into public health frameworks for cardiovascular risk prevention.^7^ However, the overwhelming majority of existing evidence addresses the relationship between air pollution and first-ever stroke occurrence. Whether ambient air pollutant exposure after an initial ischemic event influences the risk of subsequent cerebrovascular outcomes — in particular, stroke recurrence — remains largely uninvestigated.

Stroke recurrence is among the most clinically significant complications following an index ischemic event, affecting approximately 5% of survivors within the first year after discharge and conferring disproportionately high risks of cumulative neurological disability, functional dependence, and mortality compared with the initial stroke.^8–10^ Current secondary prevention strategies center on pharmacologic interventions — antiplatelet agents, anticoagulants, statins, and antihypertensives — alongside lifestyle modification.^11,12^ Notably, environmental exposures have not yet been systematically incorporated into post-stroke risk stratification frameworks. This represents a potentially important knowledge gap. After an acute ischemic event, the cerebral and systemic vasculature is characterized by heightened endothelial dysfunction, persistent inflammation, oxidative stress, and a prothrombotic state^13,14^ — conditions that may amplify the vascular toxicity of inhaled pollutants and accelerate recurrent ischemic injury. Particulate matter and gaseous pollutants can trigger systemic inflammatory responses, autonomic nervous system imbalance, and direct oxidative damage, collectively promoting atherosclerotic plaque instability and impaired cerebrovascular autoregulation.^15,16^ Despite this biological plausibility, prospective data examining whether sustained post-discharge exposure to ambient air pollutants modifies the risk of stroke recurrence are essentially absent from the current literature.

The present study was designed to address this evidence gap using data from the Clinical Research Collaboration for Stroke in Korea–National Institutes of Health (CRCS-K-NIH) multicenter prospective registry. We aimed to evaluate whether post-discharge exposure to six major ambient air pollutants — PM_10_, PM_2.5_, NO₂, SO₂, CO, and O₃ — is independently associated with the risk of stroke recurrence in patients with acute ischemic stroke.

## METHODS

### Study Population and Data Source

Data were drawn from the CRCS-K-NIH (Clinical Research Collaboration for Stroke in Korea–National Institutes of Health) registry, a nationwide, multicenter, prospective cohort that enrolls consecutive patients with acute stroke across 20 centers throughout South Korea.^17–19^ Between March 2014 and April 2021, a total of 58,370 patients were registered. The registry captures standardized clinical information including demographics, vascular risk factors, stroke characteristics, in-hospital treatment, and follow-up outcomes ascertained at scheduled intervals.

The eligibility criteria for the present analysis are detailed in Supplemental Figure 1. Patients were included if they (1) were admitted within 7 days of symptom onset, (2) had a confirmed ischemic stroke with acute infarct lesion on brain CT or MRI, (3) had provided informed consent for residential address collection, and (4) had valid ambient air quality data successfully matched to their residential district. Patients were excluded for the following reasons: missing discharge date or modified Rankin Scale (mRS) data, duplicate enrollment due to re-hospitalization, discharge mRS > 3— as patients with severe functional disability are largely confined indoors and thus have minimal exposure to ambient air pollutants, making district-level outdoor pollution concentrations a poor proxy for their actual exposure —follow-up duration less than 3 months, or occurrence of any vascular event within the first 3 months after discharge. The 3-month post-discharge exclusion window was applied to temporally separate the exposure assessment period from the outcome observation period, thereby minimizing the risk of reverse causality between early recurrence events and the assigned pollutant exposure levels. After applying all criteria, 27,346 patients constituted the final analytic cohort. The geographic distribution of patients’ residential addresses across South Korea is shown in Supplemental Figure 2.

The study protocol was approved by the institutional review boards of all participating centers, and all patients provided written informed consent. Individual patient-level data are available on reasonable request to the corresponding author, subject to legal and ethical restrictions. This study was reported in accordance with the STROBE (Strengthening the Reporting of Observational Studies in Epidemiology) reporting guidelines.^20^

### Exposure Assessment

The primary exposure was the average ambient concentration of six criteria air pollutants during the first 90 days following hospital discharge: particulate matter with aerodynamic diameter ≤10 μm (PM_10_, μg/m³) and ≤2.5 μm (PM_2.5_, μg/m³), nitrogen dioxide (NO₂, ppb), sulfur dioxide (SO₂, ppb), carbon monoxide (CO, ppb), and ozone (O₃, ppb). Hourly pollutant concentration data were obtained from the Korean National Ambient Air Monitoring Network, operated by the Ministry of Environment.

Because air quality monitoring stations are not uniformly distributed across all administrative districts (si/gun/gu), hourly pollutant and meteorological data from 332 monitoring stations were spatially interpolated to 250 districts using inverse-distance weighting (IDW).^21^ In the IDW approach, the estimated concentration at each district centroid is a weighted average of observed values from surrounding stations, with weights inversely proportional to the squared distance between the station and the target district. For each patient, district-level hourly concentrations were averaged over the 90-day post-discharge window to derive a single mean exposure value per pollutant. PM₂.₅ monitoring was not available in all districts throughout the study period, as some monitoring stations were established after the study enrollment began; consequently, PM₂.₅ exposure data were available for 26,340 of the 27,346 patients (96.3%); these 1,006 patients with missing PM₂.₅ data were excluded only from PM₂.₅ models (complete-case analysis), with no imputation applied. The distributions of post-discharge air pollutant concentrations, including median values, interquartile ranges, and histograms, are presented in Supplemental Figure 3.

Sensitivity analyses using 1-month and 2-month post-discharge averaging windows were also performed to assess the robustness of findings across different exposure durations.

### Outcome Definition

The primary outcome was stroke recurrence, defined as a new ischemic or hemorrhagic stroke event occurring more than 3 months but within 15 months after hospital discharge. This 12-month outcome observation window was defined to commence after the 3-month post-discharge exposure assessment period, thereby avoiding temporal overlap between exposure and outcome. Recurrence events were prospectively ascertained through scheduled follow-up assessments at participating centers, conducted via in-person hospital visits or structured telephone interviews by trained research coordinators.^17^ Events reported during telephone interviews were subsequently verified against medical records when available. All-cause mortality during the follow-up period was documented and treated as a competing risk in the primary analysis.

### Covariates

A comprehensive set of covariates was specified a priori and included in all multivariable models. Patient-level variables included age, sex, onset-to-arrival time, initial National Institutes of Health Stroke Scale (NIHSS) score, premorbid mRS, stroke subtype classified by the Trial of Org 10172 in Acute Stroke Treatment (TOAST) criteria using a pre-specified magnetic resonance imaging (MRI)-based algorithm,^22^ history of prior stroke or transient ischemic attack (TIA), coronary heart disease, hypertension, diabetes mellitus, dyslipidemia, atrial fibrillation, current smoking status, intravenous thrombolysis, endovascular thrombectomy, discharge prescriptions for antiplatelet agents, anticoagulants, and statins, initial fasting glucose level, and initial systolic blood pressure. District-level variables, aggregated at the si/gun/gu administrative unit, included gross regional domestic product (GRDP) per capita, marriage rate, proportion of highly educated residents, working-age population rate, green area proportion, and mean temperature, relative humidity, and atmospheric pressure during the 90-day post-discharge period. The admitting hospital was additionally included to account for center-level confounding.

### Statistical Analysis

Statistical analyses were performed using SAS version 9.4 (SAS Institute Inc., Cary, NC, USA), and graphical outputs were generated with R version 4.5.1 (R Foundation for Statistical Computing, Vienna, Austria). The primary analytic framework was a marginal Cox proportional hazards model with cause-specific hazard specification, with stroke recurrence as the event of interest and all-cause mortality as a competing risk.^23^ To account for the multilevel structure (patients nested within districts), cluster-robust standard errors were estimated. The cause-specific approach was preferred over the Fine-Gray subdistribution model to evaluate the etiological impact of exposures on recurrence while accounting for death as a competing event.^23^ Continuous exposures were modeled using restricted cubic splines to allow for nonlinear associations.

Baseline characteristics were compared across quartiles of each pollutant using the chi-squared test for categorical variables and analysis of variance or the Kruskal-Wallis test for continuous variables. Cumulative incidence functions for stroke recurrence and all-cause mortality were estimated using the Aalen-Johansen estimator and compared across quartiles of each pollutant using Gray’s test and the log-rank test, respectively. In multivariable models, each pollutant was analyzed as a categorical variable (quartiles, with the first quartile as the reference) and separately as a continuous variable. Adjusted hazard ratios (HRs) and 95% confidence intervals (CIs) were estimated from the marginal Cox model.

To evaluate potential nonlinear dose-response relationships, restricted cubic splines (RCS) with 3 knots placed at the 10th, 50th, and 90th percentiles of each pollutant distribution were fitted. Omnibus tests for the overall association (Wald test) and for departure from linearity were reported. For pollutants demonstrating a significant overall association, a threshold concentration was identified as the point at which the lower bound of the 95% confidence band first exceeded an HR of 1.0.

The proportional hazards assumption was evaluated using scaled Schoenfeld residuals and visual inspection of log-log survival plots; no substantive violations were identified.

Predefined subgroup analyses were conducted by stratifying on age (<75 vs ≥75 years), sex, stroke subtype, presence of atrial fibrillation, symptomatic steno-occlusion of the relevant artery, and mRS at discharge (0–1 vs 2–3), hypothesizing that patients with better functional status would exhibit a stronger association between ambient pollution and stroke recurrence due to their higher likelihood of outdoor exposure. Effect modification was assessed using multiplicative interaction terms between the pollutants and each stratification variable on the log-hazard scale with Cox models. All statistical tests were two-sided. While P < 0.05 was considered statistically significant for primary analyses, a more lenient threshold of P < 0.10 was applied for interaction terms. This adjustment was made to account for the conventionally reduced statistical power inherent in subgroup analyses. Given the exploratory nature of the subgroup analyses and the limited number of recurrence events, no formal correction for multiple comparisons (e.g., Bonferroni correction) was applied; interaction P values are presented as descriptive indicators of potential effect modification rather than confirmatory evidence of definitive association.

Sensitivity analyses examined the robustness of findings across 1-month, 2-month, and 3-month post-discharge exposure windows. A spatial analysis of pollutant concentration distribution across Korean administrative districts was performed to contextualize geographic patterns of high-exposure regions.

## RESULTS

Of the 58,370 patients initially registered in the CRCS-K-NIH cohort, 27,346 met all eligibility criteria and were included in the final analysis (Supplemental Figure 1). The mean age was 66.7 ± 12.9 years, and approximately two-thirds of patients were male (17,410; 63.7%). The most prevalent vascular risk factor was hypertension (63.9%), followed by diabetes mellitus (31.1%), dyslipidemia (30.4%), and atrial fibrillation (16.5%). A history of prior stroke or TIA was present in 16.1%. The most common stroke subtype by TOAST classification was large artery atherosclerosis (34.5%), followed by small vessel occlusion (23.2%) and cardioembolism (17.3%). Symptomatic steno-occlusion of the relevant artery was documented in 38.0%. The median NIHSS at admission was 2.0 (IQR 1.0–5.0), and the median mRS at discharge was 1.0 (IQR 1.0–2.0), with 55.9% of patients having mRS 0–1. At discharge, 82.4% received antiplatelet therapy, 18.5% anticoagulant therapy, and 92.5% statin therapy. Baseline characteristics are summarized in the Table and baseline characteristics stratified by quartiles of each air pollutant are provided in Supplemental Tables 1–6. Across pollutant quartiles, the distributions of most baseline variables were generally similar, although some statistically significant differences were observed for district-level meteorological and socioeconomic variables — differences that are expected given the inherent correlation between pollutant concentrations and geographic or seasonal factors.

During the follow-up period (median 364.8 days; IQR 364.8–365.8), 765 patients experienced stroke recurrence and 471 died. The cumulative incidence at 1 year was 2.80% (95% CI, 2.61–3.00) for stroke recurrence and 1.98% (95% CI, 1.82–2.15) for all-cause mortality, with each outcome treated as a competing risk for the other. Detailed cumulative incidence estimates stratified by pollutant quartiles are presented in Supplemental Table 7 for stroke recurrence and Supplemental Table 8 for all-cause mortality. Cumulative incidence functions for stroke recurrence, stratified by quartiles of each air pollutant, are presented in Figure 1. Among the six pollutants examined, Gray’s test indicated a statistically significant difference in the cumulative incidence of stroke recurrence across quartiles for SO₂ and PM_10_ (P = 0.031 and 0.034, respectively). No statistically significant separation of cumulative incidence curves was observed for NO₂, CO, O₃, or PM_2.5_. For all-cause mortality, no statistically significant differences in cumulative incidence across pollutant quartiles were observed for any of the six pollutants (all log-rank P > 0.05; Supplemental Table 8 and Supplemental Figure 4).

**Figure 1.**
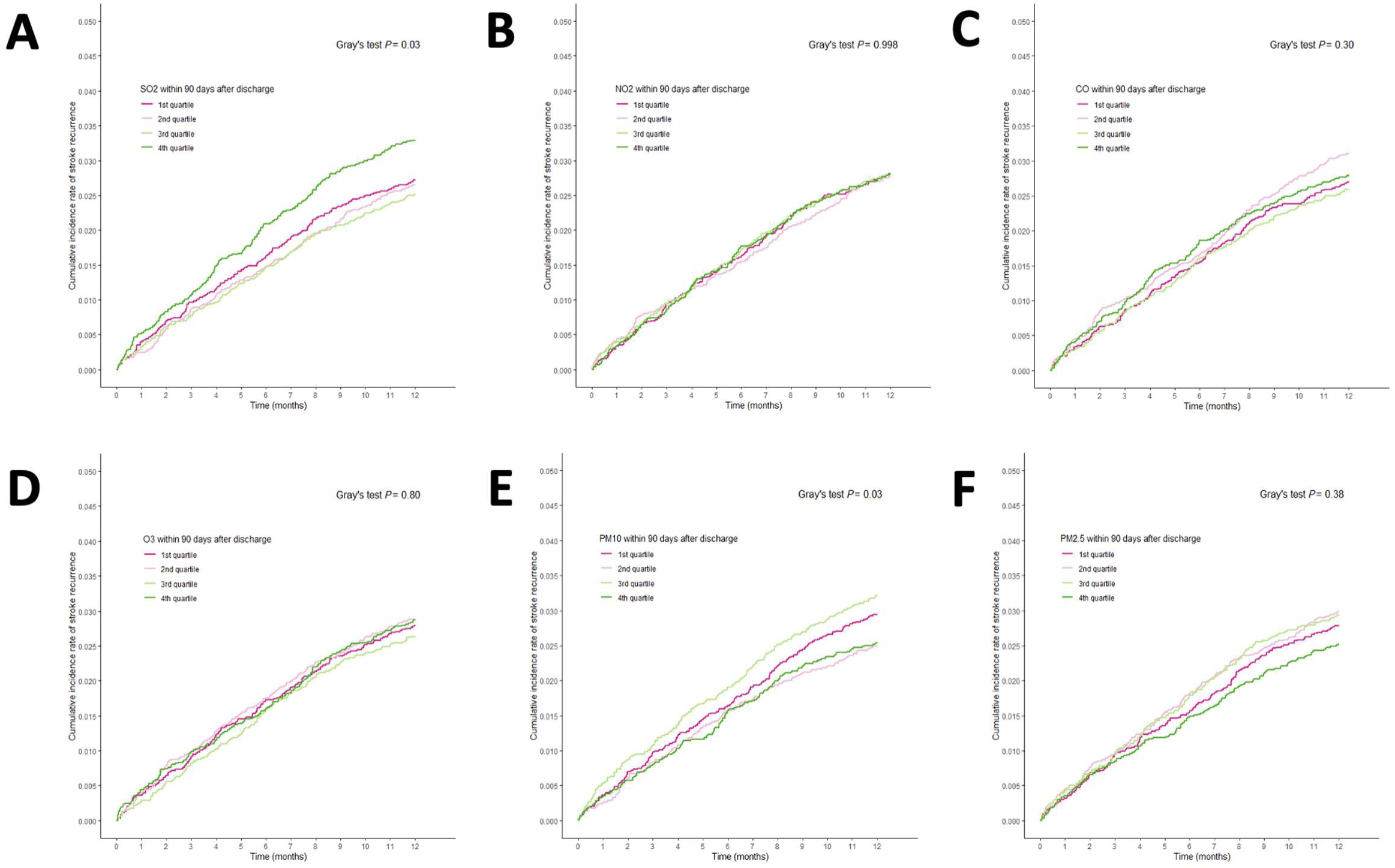
Cumulative Incidence Functions for Stroke Recurrence by Quartiles of Air Pollutant Exposure. Cumulative incidence curves for stroke recurrence within 1 year after discharge according to quartiles of the 3-month average post-discharge concentrations of **(A)** SO₂, **(B)** NO₂, **(C)** CO, **(D)** O₃, **(E)** PM10, and **(F)** PM2.5. All-cause mortality was treated as a competing risk. *P* values were derived using Gray’s test.

In the multivariable marginal Cox model, SO₂ was the only pollutant demonstrating a statistically significant overall association with stroke recurrence (P for overall effect = 0.049; Supplemental Table 9). Compared with the lowest quartile, the adjusted hazard ratios for the second, third, and fourth SO₂ quartiles were 0.96 (95% CI, 0.78–1.19), 0.91 (95% CI, 0.73–1.13), and 1.19 (95% CI, 0.94–1.50), respectively. While individual pairwise quartile comparisons did not reach statistical significance, the omnibus test across all quartiles was significant, with increased risk concentrated in the highest exposure category. The remaining five pollutants showed no significant overall associations (Supplemental Table 9).

Restricted cubic spline analysis modeling SO₂ as a continuous variable revealed a J-shaped dose-response curve, with the hazard of stroke recurrence remaining relatively flat at lower concentrations and rising progressively at higher exposure levels (Figure 2). The omnibus Wald test for the overall association in this spline model was statistically significant (P = 0.024), while the test for departure from linearity approached significance (P = 0.095), suggesting a potential threshold effect. The concentration at which the lower bound of the 95% confidence interval first exceeded an HR of 1.0 was identified at approximately 8.2 ppb. As this threshold was identified post hoc from the spline curve and the test for non-linearity did not reach conventional statistical significance, the threshold should be regarded as exploratory and requires confirmation in independent cohorts. For the remaining five pollutants, RCS analyses demonstrated no significant overall associations, and the spline curves were generally flat without discernible inflection points (Figure 2).

**Figure 2.**
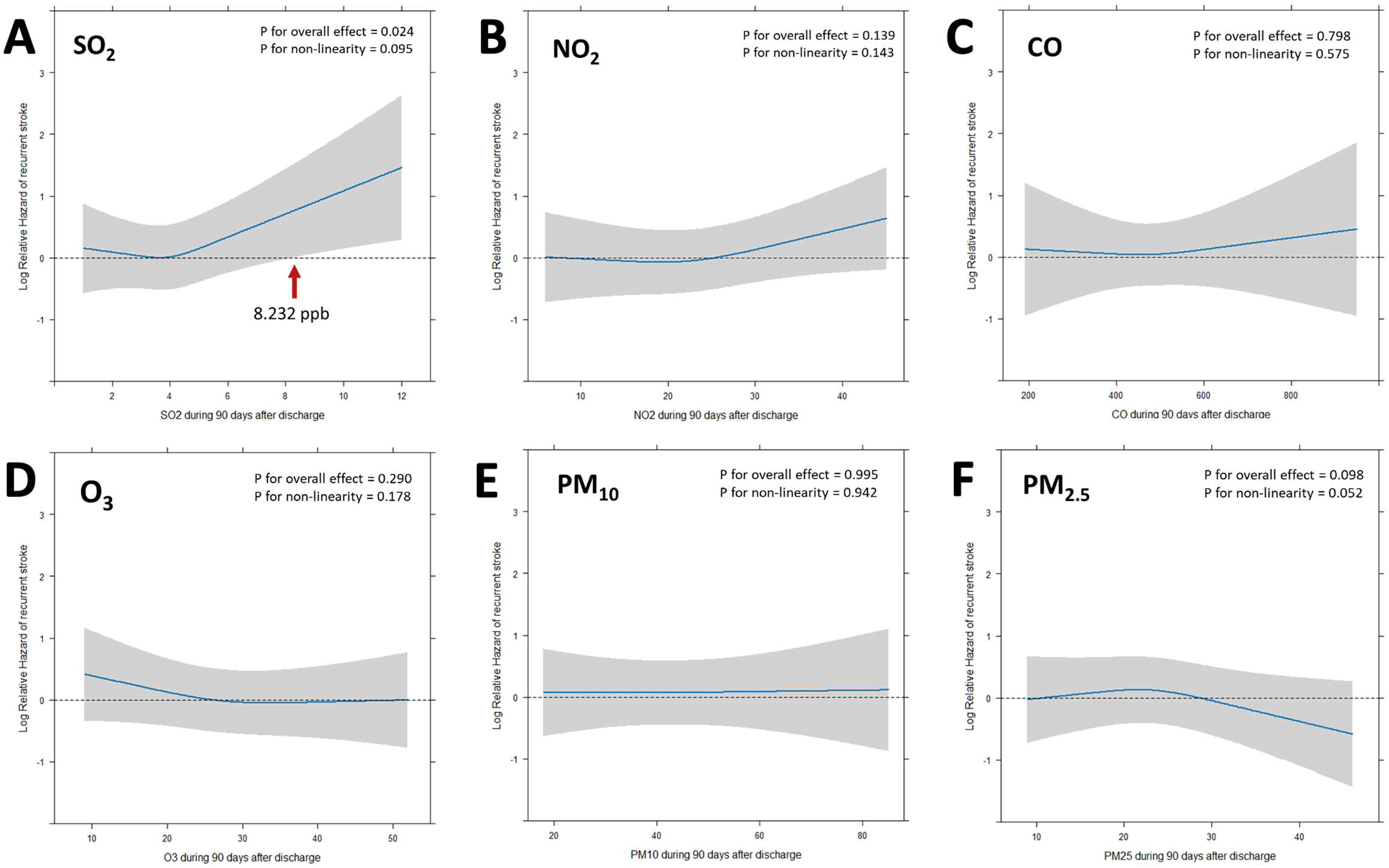
Restricted Cubic Spline Plots for Air Pollutants and Stroke Recurrence. Restricted cubic spline curves showing the association between each air pollutant, modeled continuously, and the cause-specific hazard of stroke recurrence: **(A)** SO₂, **(B)** NO₂, **(C)** CO, **(D)** O₃, **(E)** PM10, and **(F)** PM2.5. The reference value was the median exposure level. SO₂, NO₂, O₃, and CO are expressed in ppb; PM10 and PM2.5 are expressed in μg/m³. The horizontal dashed line indicates HR = 1.0. In panel A, the SO₂ threshold of 8.232 ppb is marked, corresponding to the point where the upper 95% CI first exceeded HR = 1.0. *P* values for overall association and nonlinearity are shown in each panel.

Predefined subgroup analyses for SO₂ are summarized in Supplemental Figure 5. In restricted cubic spline models stratified by predefined subgroups, the dose-response association between SO₂ exposure and stroke recurrence was numerically stronger among older adults aged ≥75 years (P for interaction = 0.051) and among women (P for interaction = 0.062), although both interaction terms approached but did not reach conventional statistical significance. No meaningful effect modification was observed by stroke subtype, atrial fibrillation status, symptomatic steno-occlusion, or discharge mRS.

Spatial analysis revealed substantial geographic heterogeneity in SO₂ concentrations across Korean districts (Figure 3). The highest average SO₂ levels during the study period were concentrated in major port cities: Incheon Metropolitan City Jung-gu (9.5 ppb), Ulsan Metropolitan City Nam-gu (8.9 ppb), Busan Metropolitan City Yeongdo-gu (7.3 ppb), Ulsan Metropolitan City Dong-gu (7.2 ppb), and Busan Metropolitan City Jung-gu (7.1 ppb). Notably, two of these five districts exceeded the exploratory recurrence threshold of approximately 8.2 ppb. This geographic distribution is consistent with known sources of SO₂ emissions from maritime shipping and petrochemical industrial activities.

**Figure 3.**
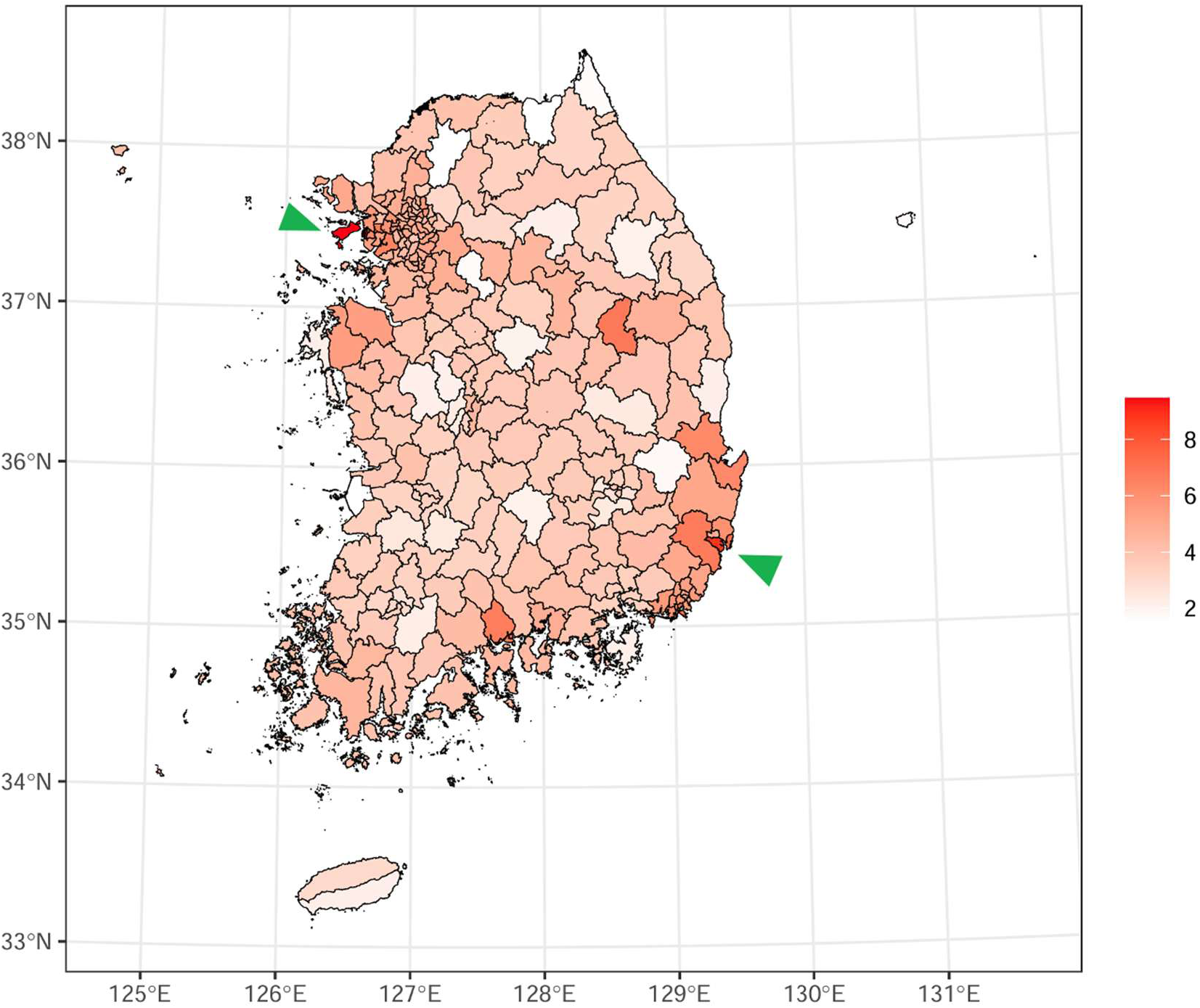
Spatial Distribution of Average SO₂ Concentration Across Korean Districts (2014–2021). Geographic map of South Korea depicting the average ambient SO₂ concentration by district (si/gun/gu) over the study period. The identified SO₂ recurrence threshold of 8.232 ppb is indicated. Two districts above the SO₂ threshold level — Incheon Jung-gu (9.5 ppb) and Ulsan Nam-gu (8.9 ppb) — are annotated with green arrowheads, highlighting the concentration of high SO₂ exposure in major port cities.

Sensitivity analyses using shorter post-discharge exposure windows revealed a pattern consistent with a cumulative dose-response relationship (Figure 4). When SO₂ exposure was averaged over the 1-month post-discharge period, the association with stroke recurrence did not reach statistical significance; however, the P value decreased with the 2-month exposure window, and the association became statistically significant with the 3-month window. This progressive strengthening of the association with longer exposure durations suggests that the cerebrovascular effect of SO₂ may be cumulative, requiring sustained exposure above a threshold concentration to appreciably elevate recurrence risk. The J-shaped dose-response pattern was preserved across all three exposure windows.

**Figure 4.**
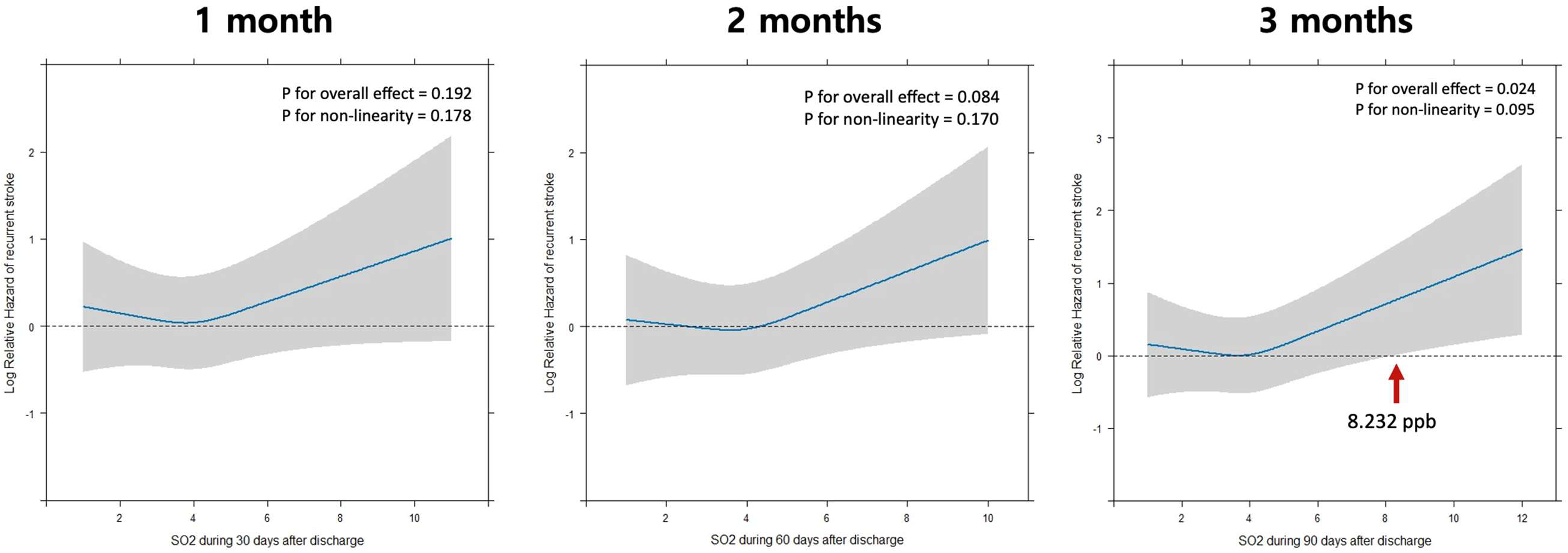
Sensitivity Analysis: SO₂ Dose-Response by Exposure Duration. Restricted cubic spline plots for SO₂ as a continuous variable, repeated for 1-month, 2-month, and 3-month post-discharge exposure windows. SO₂ is expressed in ppb. The association did not reach statistical significance with the 1-month window but progressively strengthened with longer exposure durations, achieving significance at 3 months. This pattern suggests a cumulative dose-response relationship, whereby sustained SO₂ exposure above a threshold concentration is required to appreciably elevate recurrence risk. The J-shaped dose-response pattern was preserved across all three windows.

## DISCUSSION

In this nationwide multicenter registry study of 27,346 patients with acute ischemic stroke, we found that among six major ambient air pollutants assessed during the first 3 months after hospital discharge, only sulfur dioxide demonstrated a statistically significant association with stroke recurrence within one year. A potential exposure threshold was identified at approximately 8.2 ppb, above which the risk of recurrence increased progressively. This association appeared more pronounced among older adults and women, and the highest SO₂ concentrations were geographically concentrated in major port cities. To our knowledge, this is the first study to evaluate the association between post-discharge ambient air pollutant exposure and stroke recurrence using a competing risk framework in a large multicenter registry cohort.

The observation that SO₂ — rather than particulate matter or other gaseous pollutants — was the sole pollutant significantly associated with stroke recurrence merits careful interpretation. SO₂ has distinct pathophysiologic properties relevant to cerebrovascular injury. As a highly water-soluble gas, SO₂ is absorbed by the respiratory tract and subsequently enters the systemic circulation in the form of sulfite and bisulfite derivatives.^24,25^ Experimental evidence in rat models of middle cerebral artery occlusion has demonstrated that sustained SO₂ inhalation upregulates cortical expression of endothelin-1, inducible nitric oxide synthase, cyclooxygenase-2, and intercellular adhesion molecule-1, thereby promoting endothelial dysfunction, vascular inflammation, and exacerbation of ischemic brain injury.^24^ These pathways are particularly relevant in the post-stroke vasculature, which is already characterized by endothelial injury, heightened inflammatory tone, and a prothrombotic milieu following the index event.

Epidemiologic data are consistent with these mechanistic observations. Multiple studies have shown that both short- and long-term exposure to SO₂ is associated with increased hospital admissions for cardiovascular disease and ischemic stroke.^5,26,27^ A Korean nationwide study reported a particularly strong association between SO₂ and cardioembolic stroke (adjusted OR 1.57 per 10-ppb increment), raising the possibility that SO₂ exposure may trigger atrial fibrillation — a well-established mechanism of cardioembolic stroke — through autonomic dysregulation, atrial oxidative stress, and inflammatory remodeling of the atrial myocardium.^28^ Indeed, ambient air pollution has been linked to increased risk of new-onset and paroxysmal atrial fibrillation in several population-based studies, providing a plausible pathway through which SO₂ could preferentially elevate cardioembolic stroke risk.^27,29^

The geographic concentration of high SO₂ exposure in harbor regions creates a setting in which a subpopulation of patients experiences persistently elevated exposure from maritime shipping emissions.^30,31^ While maritime shipping also emits PM2.5 and NOx, urban concentrations of these pollutants are substantially influenced by non-maritime sources, including traffic and residential combustion, which may dilute the harbor–non-harbor exposure contrast.^32^ The relatively pure source-specificity of SO₂ may therefore yield greater spatial heterogeneity, enhancing statistical power to detect an association.

The absence of significant associations for the other five pollutants should not be interpreted as definitive evidence of no effect. The relatively modest number of recurrence events (n = 765) may have limited statistical power, particularly for pollutants with narrower interquartile ranges. Notably, PM₂.₅ showed a borderline overall association (P = 0.098) with a suggestion of non-linearity (P = 0.052); however, the direction of the association was inverse — with higher concentrations associated with lower recurrence risk — which is counterintuitive from a biological standpoint. This paradoxical finding likely reflects residual confounding by urbanization-related factors: in South Korea, PM₂.₅ concentrations tend to be highest in densely populated metropolitan areas,^33,34^ where residents also have greater access to specialized stroke care, shorter hospital transport times, higher socioeconomic status, and better medication adherence — all of which independently reduce recurrence risk.^35–37^ Although we adjusted for district-level socioeconomic indicators, these covariates may not have fully captured the protective health infrastructure advantages associated with urban residence.

The spatial analysis revealed that the highest SO₂ concentrations were concentrated in Incheon, Ulsan, and Busan — South Korea’s principal maritime shipping and petrochemical industrial hubs. The International Maritime Organization (IMO) implemented a global sulfur cap in January 2020, reducing the permissible sulfur content of marine fuels from 3.5% to 0.5%, which has been associated with substantial reductions in SO₂ emissions in designated emission control areas worldwide.^38,39^ However, our study period (2014–2021) largely predates the full implementation of these regulations, and the observed SO₂ levels in harbor districts may reflect a transitional pollution environment. Whether the post-2020 decline in shipping-related SO₂ emissions has translated into reduced cerebrovascular risk among harbor-region residents remains an important question for ongoing surveillance. SO₂ also remains a major concern in regions where fossil fuel combustion continues to be a dominant energy source, where ambient levels substantially exceed those observed in Korea.^40,41^

The tendency for a stronger SO₂–recurrence association in older adults and women is biologically plausible in the post-stroke context. Aging is accompanied by inadequate antioxidant defenses, with insufficient upregulation of superoxide dismutase, catalase, and glutathione in the face of increased reactive oxygen species, as well as endothelial dysfunction characterized by reduced nitric oxide bioavailability and impaired endothelium-dependent vasodilation,^42^ which may lower the threshold at which exogenous oxidative stressors are sufficient to provoke recurrent vascular injury. Prior studies have reported that older individuals exhibit heightened cardiovascular susceptibility to ambient air pollution.^43^

With respect to sex differences, the loss of estrogen-mediated vascular protective effects in postmenopausal women may reduce nitric oxide bioavailability and diminish anti-inflammatory signaling, thereby heightening susceptibility to pollution-induced endothelial activation.^44–46^ Epidemiologic evidence has indicated that women may be more susceptible to the adverse cardiovascular effects of air pollution than men.^47,48^ Despite recent improvements in SO₂ regulation, these high-risk populations may remain particularly vulnerable, underscoring the need for ongoing monitoring and regional pollution control. These subgroup findings, however, should be considered hypothesis-generating and require confirmation in dedicated analyses with adequate statistical power.

The clinical implications of our findings are twofold. At the individual patient level, the identification of a potential SO₂ threshold (approximately 8.2 ppb) provides an approximate reference concentration that could inform risk communication during discharge planning. For patients residing in high-SO₂ districts, clinicians might consider advising awareness of local air quality indices, recommending limitation of outdoor activity on days of elevated SO₂ levels, and ensuring optimization of evidence-based secondary prevention medications, which may partially mitigate pollution-related vascular injury. At the population level, these findings strengthen the rationale for targeted air quality monitoring and intervention not only in communities adjacent to major ports but also in regions where fossil fuel combustion remains a predominant source of SO₂ emissions. The integration of ambient SO₂ surveillance data with stroke registry systems could enable real-time identification of high-risk geographic areas and inform regionally tailored prevention strategies. More broadly, these results support the emerging concept that secondary stroke prevention should extend beyond individual-level pharmacotherapy to encompass the environmental context in which patients live after discharge.^7^

This study has several notable strengths. The CRCS-K-NIH registry provided a large, nationwide, multicenter cohort with standardized data collection, prospective outcome ascertainment, and comprehensive covariate information across 20 centers, enhancing both generalizability and the ability to control for confounding.^8^ The use of a cause-specific hazard model within a marginal Cox framework appropriately accounts for the competing risk of death.^23^ The inclusion of district-level socioeconomic and meteorological covariates addresses confounding pathways often overlooked in air pollution studies.^49^ The consistency of findings across multiple sensitivity analyses using different exposure windows supports the robustness of the observed association.

Several limitations should be acknowledged. First, as an observational study, residual confounding cannot be entirely excluded despite adjustment for an extensive covariate set. Unmeasured factors such as indoor air pollution exposure, residential proximity to specific emission sources, and individual-level activity patterns may have influenced the results. Second, exposure was assigned at the district level using spatially interpolated data rather than personal monitoring — an approach necessitated by the scale of the study — which likely introduces non-differential measurement error and may attenuate true effect estimates toward the null.^50^ Third, we did not account for residential mobility during the post-discharge period; patients who moved to a different district would have been misclassified with respect to their actual pollution exposure. Fourth, the single-pollutant modeling approach does not capture potential synergistic or antagonistic interactions among co-occurring pollutants. Relatedly, the observed association with SO₂ may not reflect a direct causal effect of SO₂ itself but could instead serve as a proxy for co-emitted pollutants that were not independently measured in this study — such as ultrafine particles, volatile organic compounds, or other combustion-related byproducts that share common emission sources with SO₂, particularly maritime shipping and industrial activities.^51^ Fifth, outcome ascertainment through clinical follow-up may not have captured all recurrent events, particularly mild strokes managed at non-participating hospitals, which could lead to underestimation of recurrence rates. Sixth, the study period (2014–2021) overlaps with the COVID-19 pandemic, during which both ambient air pollution levels and healthcare-seeking behavior changed substantially; lockdown-related reductions in pollutant emissions and delays in stroke presentation may have affected 2020–2021 data.^52–54^

## CONCLUSIONS

In this nationwide multicenter registry study of acute ischemic stroke patients, post-discharge exposure to ambient sulfur dioxide was associated with an increased risk of stroke recurrence within one year. A potential threshold effect was identified at approximately 8.2 ppb, with the association being more pronounced in older adults and women. The highest SO₂ concentrations were geographically concentrated in harbor regions with active maritime shipping. These findings suggest that ambient SO₂ may represent a modifiable environmental determinant of stroke recurrence risk and support the integration of air quality considerations into secondary stroke prevention strategies. Future studies should confirm these findings in independent cohorts and evaluate whether reductions in shipping-related SO₂ emissions translate into lower recurrence rates in exposed populations.

### Source of funding

This research was supported by Basic Science Research Program through the National Research Foundation of Korea (NRF) funded by the Ministry of Education (grant number: RS-2023-00238058) and a grant of the “Korea National Institute of Health” research project (project No. 2023-ER-1006-02). The funding sources did not participate in any part of the study, from conception to article preparation.

### Disclosures

Hee-Joon Bae reports grants from Amgen Korea Inc., Bayer Korea, Bristol Myers Squibb Korea, Celltrion, Dong-A ST, Otsuka Korea, Samjin Pharm, and Takeda Pharmaceuticals Korea Co., Ltd., and personal fees from Amgen Korea, Bayer, Daewoong Pharmaceutical Co., Ltd., Daiichi Sankyo, Esai Korea, Inc., JW Pharmaceutical, and SK chemicals, outside the submitted work.

## Supplemental Material

Supplemental Tables 1 to 9

Supplemental Figures 1 to 5

STROBE checklist

## Data Availability

Individual patient-level data are available on reasonable request to the corresponding author, subject to legal and ethical restrictions.

## Non-standard Abbreviations and Acronyms

CO: carbon monoxide
COVID-19: coronavirus disease 2019
CRCS-K-NIH: Clinical Research Collaboration for Stroke in Korea–National Institutes of Health
GRDP: gross regional domestic product
IDW: inverse-distance weighting
IMO: International Maritime Organization
mRS: modified Rankin Scale
NIHSS: National Institutes of Health Stroke Scale
NO₂: nitrogen dioxide
O₃: ozone
PM₁₀: particulate matter with aerodynamic diameter ≤10 μm
PM₂.₅: particulate matter with aerodynamic diameter ≤2.5 μm
RCS: restricted cubic splines
SO₂: sulfur dioxide
STROBE: Strengthening the Reporting of Observational Studies in Epidemiology
TIA: transient ischemic attack
TOAST: Trial of Org 10172 in Acute Stroke Treatment

---

**Table.**
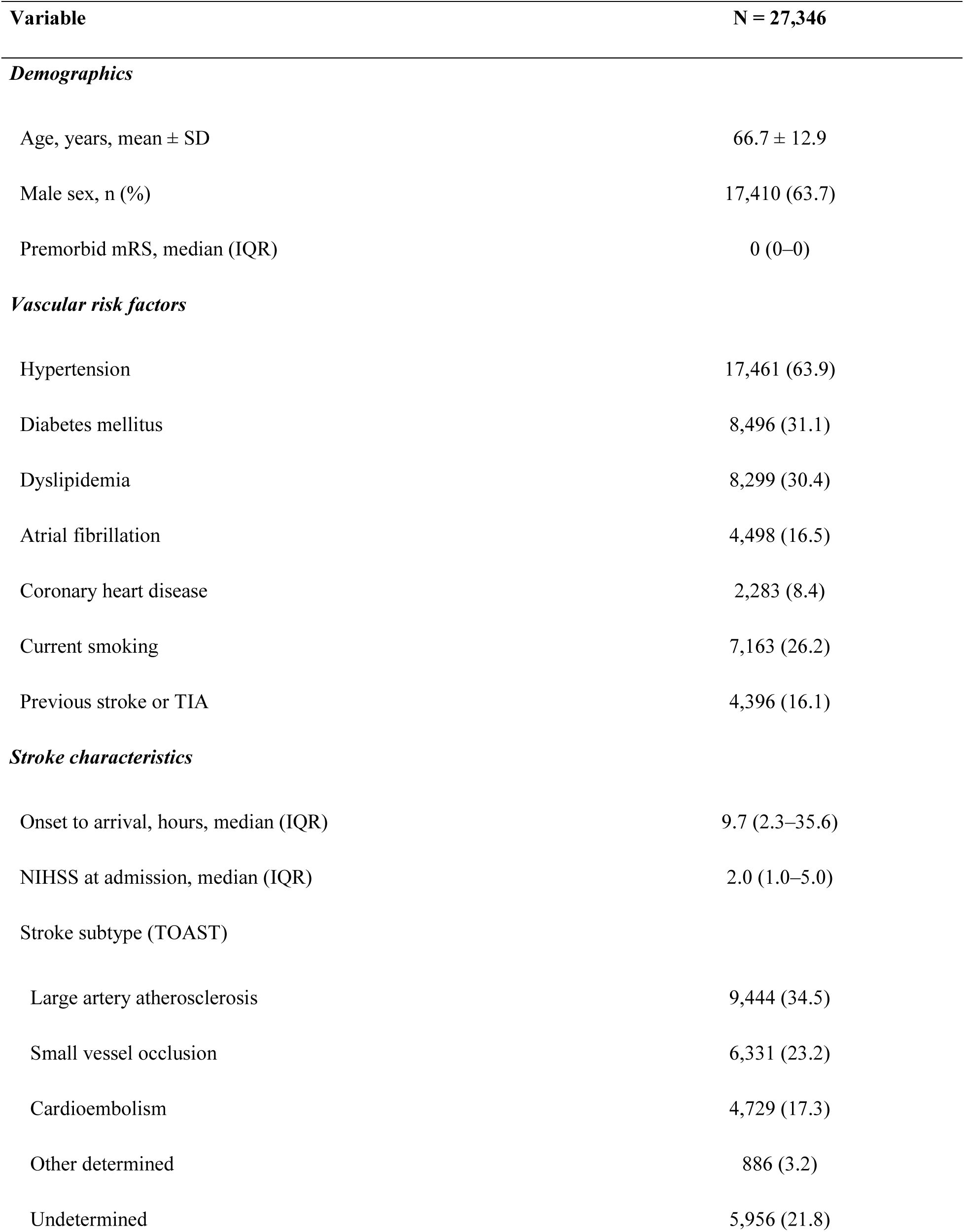
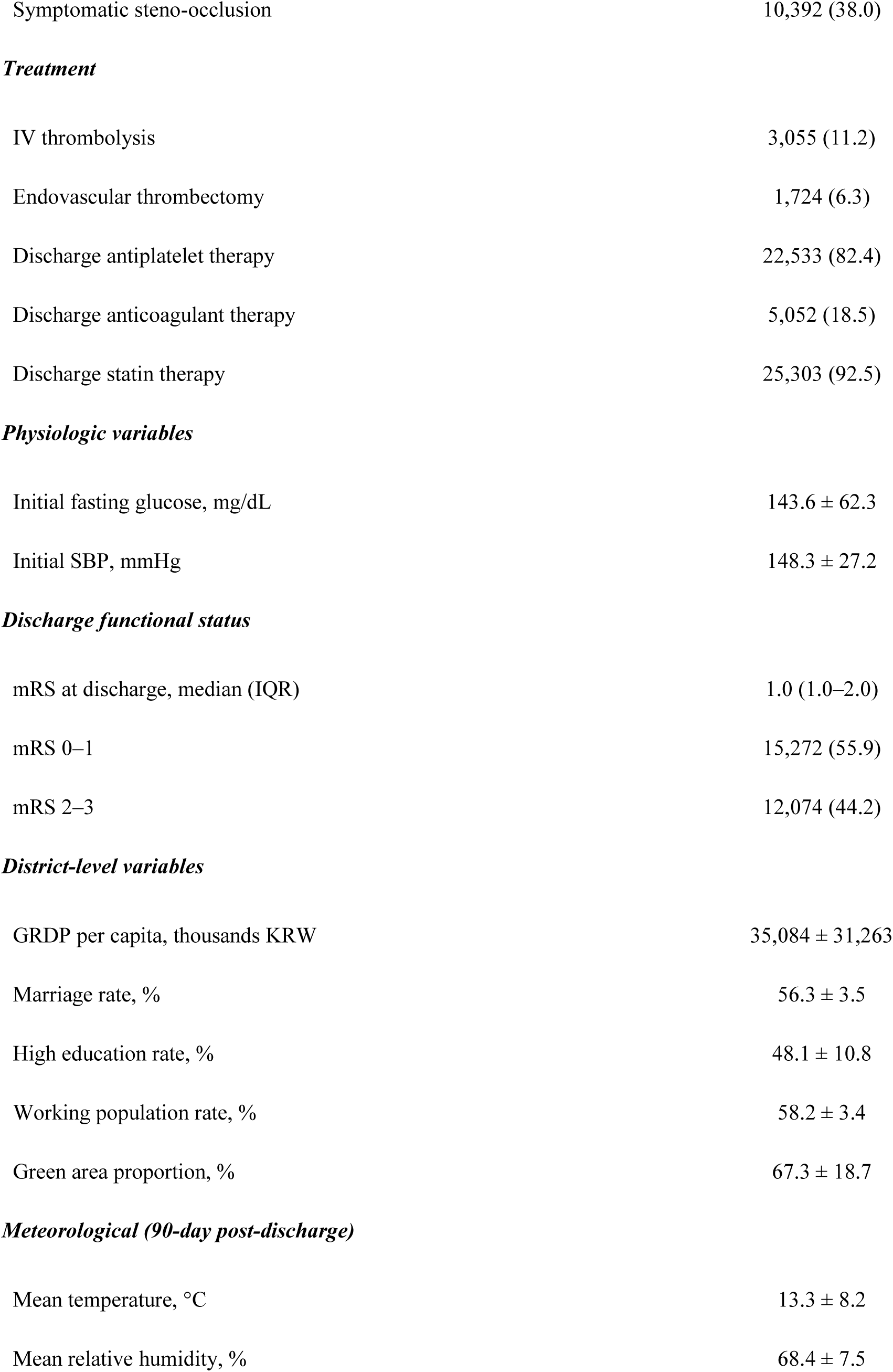
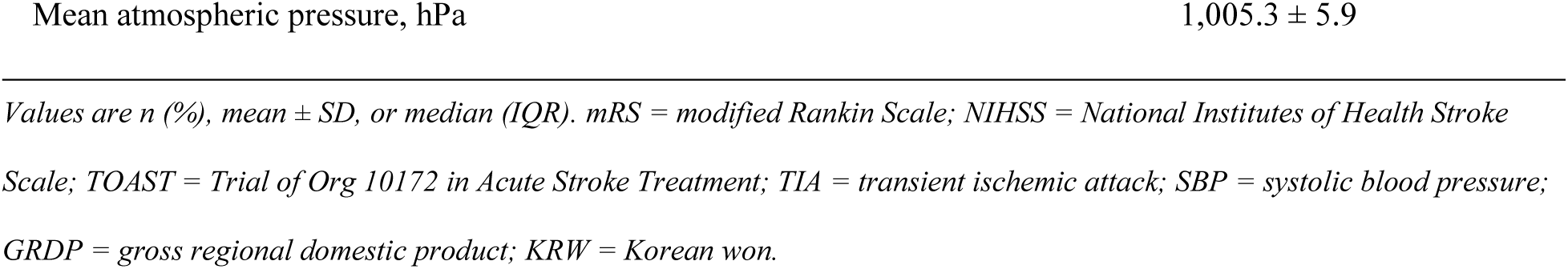
Baseline Characteristics of the Study Cohort.

